# The effects of a provincial opioid prescribing standard on opioid prescribing for pain: interrupted series analysis

**DOI:** 10.1101/2025.03.14.25323994

**Authors:** Dimitra Panagiotoglou, Sandra Peterson, M Ruth Lavergne, Tara Gomes, Rashmi Chadha, Philippa Hawley, Rita McCracken

**Affiliations:** Department of Epidemiology, Biostatistics and Occupational Health, McGill University, Montreal, Quebec, Canada; Centre for Health Services and Policy Research, University of British Columbia, Vancouver, British Columbia, Canada; Department of Family Medicine, Faculty of Medicine, Dalhousie University, Halifax, Nova Scotia, Canada; Li Ka Shing Knowledge Institute, Unity Health, Toronto, Ontario, Canada; Leslie Dan Faculty of Pharmacy, University of Toronto, Toronto, Ontario, Canada; Vancouver Coastal Health Authority, Vancouver, British Columbia, Canada; Department of Family Practice, University of British Columbia, Vancouver, British Columbia, Canada; BC Cancer Vancouver Centre and Division of Palliative Medicine, Dept. Medicine, University of British Columbia

## Abstract

**Background:** In 2016, the College of Physicians and Surgeons of British Columbia released a legally enforceable opioid prescribing practice standard for the treatment of chronic non-cancer pain (CNCP). The standard was revised in 2018, following physicians, patient groups and key partners’ concerns it was inappropriately interpreted. We tested the effects of the practice standard on access to opioids for people living with CNCP; and spillover effects on people living with cancer or receiving palliative care.

**Methods:** We used comprehensive administrative health data and multiple baseline interrupted time series analysis to evaluate the effects of the 2016 practice standard and 2018 revision.

**Results:** The practice standard accelerated pre-existing declining trends in morphine milligram equivalents (MME) dispensed per person living with CNCP (−0.1%, 95% CI: -0.2, 0.0%), but also for people living with cancer (−0.7%, 95% CI: -1.0, -0.5%) or receiving palliative care (−0.3%, 95% CI: -0.5, 0.0%). Trends for the proportion of people with CNCP prescribed an opioid >90 MME daily dose (−0.3%, 95% CI: -0.4, 0.2%), co-prescribed benzodiazepine or other hypnotic (−0.6%, 95% CI: -0.7, -0.5%), and rapidly tapered (0.1%, 95% CI: -0.2, 0.0%) also declined more quickly. While level effects were generally in the same direction, the proportion of people rapidly tapered immediately post-implementation increased 2.0% (95% CI: 0.4, 3.3%). Trends slowed or reversed post-2018 revision.

**Interpretation:** The 2016 practice standard was associated with an immediate and long-lasting effect on physicians’ opioid prescribing behaviours, including negative spillover effects on tapering, and for people living with cancer or receiving palliative care.

## INTRODUCTION

Between 2014 and 2023 there were approximately 14,975 drug deaths in British Columbia, with opioids present in over 80% of events.(1, 2) As part of the effort to lower the risk of opioid poisoning, the College of Physicians and Surgeons of British Columbia (henceforth ‘College’) released the *Safe Prescribing of Drugs with Potential for Misuse/Diversion* practice standard in June 2016 (Box 1)(3). This legally enforceable standard was developed to “prevent an increasing toll of prescription drug misuse and overdose deaths” by limiting access to opioid prescriptions for chronic noncancer pain (CNCP) management.(3) Previous treatment guidelines within Canada and abroad (e.g., CDC Guideline for Prescribing Opioids for Chronic Pain – United States (2016), the College’s *Prescribing Principles* (2012), and the National Opioid Use Guideline Group’s *Safe and Effective Use of Opioids for Chronic Non-Cancer Pain* (2010))(4, 5) recommended courses of action and allowed physicians to “exercise reasonable discretion in their decision to act on guidance provided”.(6) Conversely, the 2016 practice standard required specific prescribing practices. Physicians found noncompliant could be disciplined or fined under the Health Professions Act, RSBC 1996, c.183 and College Bylaws.(6, 7)

Across Canada and the United States (US), the effect of prescribing guidelines on access to opioids appears mixed. A study on the College’s 2016 practice standard found no change in total opioid consumption, but observed a decline in prescription renewals.(8) However, the study was not specific to CNCP because it included opioids dispensed to exempt patients (i.e., cancer or palliative care). Similarly, three US studies on opioid prescribing post-guideline implementation reported an increase in prescribing prevalence,(9-11) while two found decreased prescribing incidence,(12, 13) and two reported lower dosages(14, 15). Notably, these studies also did not focus on CNCP. Additionally, evidence suggests misinterpretation of guidelines led to non-consensual aggressive tapering, debilitating pain, and increased overdose risk.(16-22) In June 2018, the College released the “*Safe Prescribing of Opioids and Sedatives*” revision to address physicians’, patient groups’, and key partners’ concerns about misinterpretations of the 2016 standard.(23) Changes included softer language on co-prescribing opioids with sedatives, removal of stimulants from the standard, and elimination of the limit on supply.

We tested the effects of the 2016 practice standard and its 2018 revision on the morphine milligram equivalents (MME) dispensed to patients with CNCP managed with long-term opioid treatment (LTOT), and the proportion of these patients with: a) a reassessment by their prescribing physician every three months, b) a co-occurring benzodiazepine or other sedative hypnotic prescription, c) prescriptions exceeding a three-month supply or 250 pills, d) a daily dose exceeding 90 MME, or e) an opioid taper exceeding 10%. We also tested the effects of the practice standard and its revision on MME dispensed to patients living with cancer or receiving palliative care.

## METHODS

This study used data on all opioid prescriptions dispensed to non-institutionalized British Columbians between 1 October 2012 and 31 March 2020.

### Study design

We used a multiple intervention time series study design to test the effects of the practice standard and its revision. Interrupted time series analysis is a quasi-experimental study design that estimates population-level effects of policy interventions before and after implementation in contexts where randomized controlled trials are not feasible or ethical.(24) This before-after comparison assumes selection bias and within-group characteristics that change slowly over time, secular changes, random fluctuations from one time point to the next, and regression to the mean are sufficiently controlled.

### Data

We linked the BC Ministry of Health’s PharmaNet,(25) Discharge Abstract Database (DAD),(26) Medical Services Program (MSP) billings and consolidation files,(27)and National Ambulatory Care Reporting System (NACRS),(28) with the BC Cancer Agency’s cancer registry(29) and Vital Statistics’ Mortality(30) data at the patient-level to determine the primary purpose of each opioid prescription dispensed, and when to censor patients. PharmaNet included all prescriptions filled by community pharmacies to BC residents, irrespective of payer. DAD, MSP and NACRS data captured hospital and outpatient physician visits. The consolidation file and mortality datasets informed when patients first entered and exited the cohort. The cancer registry included all residents diagnosed with cancer. The individual-level, de-identified data were provided by Population Data BC. Access to the data is subject to approval, but can be requested for research projects through the Data Steward(s) or their designated providers. All inferences, opinions, and conclusions drawn in this publication are those of the authors, and do not reflect the opinions or policies of the Data Steward(s). Readers can find further information regarding these data sets by visiting the Population Data BC project webpage at: https://my.popdata.bc.ca/project_listings/20-165/

### Cohort

Our primary cohort included all adult residents (≥18 years of age) living in the community with CNCP on LTOT. To create the cohort, we identified all unique opioid prescriptions dispensed between October 2012 and March 2020 (n=46,629,134, Figure 1). We excluded opioid formulations prescribed for the management of opioid use disorder, cough or diarrhea; prescriptions dispensed after patients met cancer, palliative care or long-term residence exclusion criteria or death/end of registration; medications within seven days of trauma or major surgical intervention; prescriptions dispensed by a non-BC physician (e.g., nurse practitioners, out of province physicians, etc.) or missing the prescribing physician’s identifier. Using the remaining prescriptions, we defined people on LTOT as those with an opioid analgesic dispensation renewed at least once with at least 60 days’ supply dispensed during a 90-day period (n=9,611,066 dispensations for primary analysis retained). We repeated the process to create our spillover cohorts, retaining 870,834 prescriptions dispensed to patients living with cancer and 744,725 prescriptions to patients receiving palliative care.

**Figure 1.**
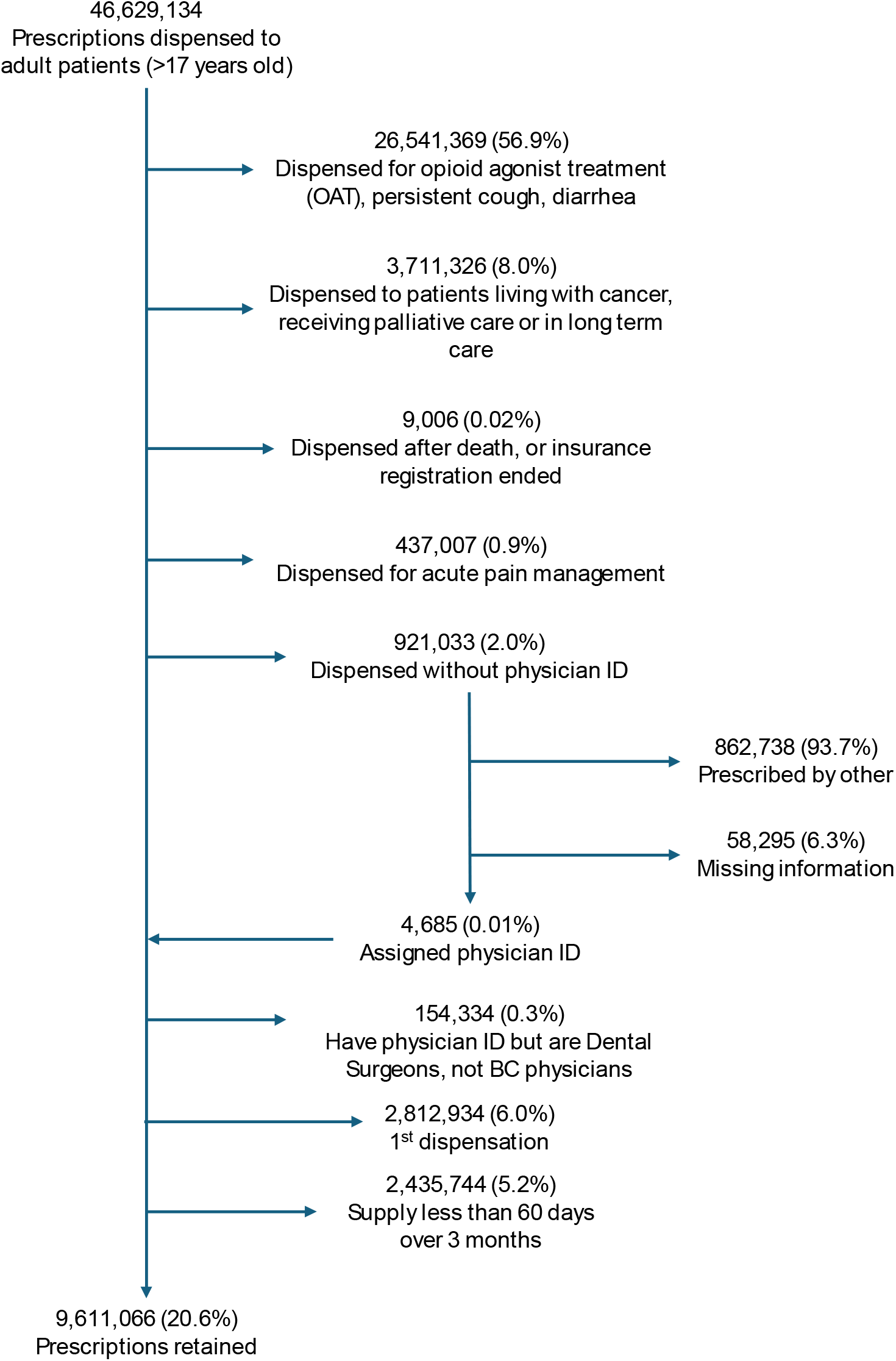
Flowchart of opioid prescriptions captured in PharmaNet dataset and retained to identify cohort of patients treated for chronic non-cancer pain dispensed long-term opioid treatment

### Ethics approval

This study was approved by McGill University’s Institutional Review Board (no. A11-M55-19A).

### Outcome

Our unit of analysis was person-month. We converted the total dose of opioids dispensed across prescriptions to daily MME using each prescription’s start date, days supply and dispensed quantity (allowing for multiple co-occurring prescriptions). We tested the effect of the practice standard on MME dispensed per patient treated with LTOT for CNCP, living with cancer or receiving palliative care, separately. We also examined the proportion of patients on LTOT who:

1. Did not visit the opioid-prescribing physician within 90 days after the prescription was dispensed;
2. Had co-occurring benzodiazepine or other hypnotic prescription at time of fill; or
3. Had a prescription that exceeded:
  a. three-month supply or 250 pills (whichever was less);
  b. daily dose of 90 MME; or
  c. dose reduction of 10% from the previous 2-week period (including abrupt cessation not due to moving out-of-province, death, being diagnosed with cancer or starting palliative care).

### Statistical analysis

We divided the study into three periods: pre-intervention (January 2013 to June 2016), post-2016 practice standard (July 2016 to June 2018), and post-2018 practice standard revision (July 2018 – December 2019). Prescription data from October 2011 – December 2012, and January – March 2020 served as wash-in and wash-out periods for our measures, respectively.

We tested the effects of the practice standard using generalized linear regression models (Poisson) for interventions j and k:

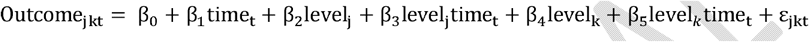

Where β_0_ was the expected outcome at the first time point (i.e., intercept); β_1_ was the pre-existing trend in the outcome; β_2_ was the change in the outcome level between the pre-intervention period and immediately following the practice standard’s implementation (June 2016); β_3_ was the change in outcome trend from the pre-intervention period to after the implementation of the 2016 practice standard; β_4_ was the change in the outcome level between the post-2016 practice standard period and its revision in June 2018; and β_5_ was the change in the outcome trend between the post-2016 practice standard period and its 2018 revision. We used the two-sided Durbin-Watson test and residual plots to test for autocorrelation (e.g. first order) and moving averages to inform the models’ correlation structures. We used Newey-West standard errors with a lag of 3 to correct for the autocorrelation detected. The data was prepared using SAS 9.4, and analysis was executed using R version 4.0.

## RESULTS

During the observation period, there were between 38,796 (February 2019) and 57,276 (October 2014) British Columbians with CNCP included in our study. We also had between 2652 (February 2013) and 7945 (October 2018) patients living with cancer, and between 1840 (November 2014) and 2231 (October 2019) patients receiving palliative care, after applying all our exclusions. At the start of the observation period (January 2013), among patients receiving LTOT for CNCP, just under 26% did not have a visit with the prescribing physician within 90 days, 29.3% had a co-occurring benzodiazepine or other hypnotic prescription dispensed, 5.7% had a supply that exceeded 90 days or 250 pills, 32.7% had a daily dose that exceeded 90 MME, and 39.2% were tapered rapidly.

Our analysis revealed that for CNCP patients, the 2016 practice standard accelerated pre-existing declining trends in the MME dispensed per person by 0.1% (95% CI: -0.2 – 0.0%) (Table 1, Figure 2), and in the proportion of patients (Table 2, Figure 3):

**Table 1.**
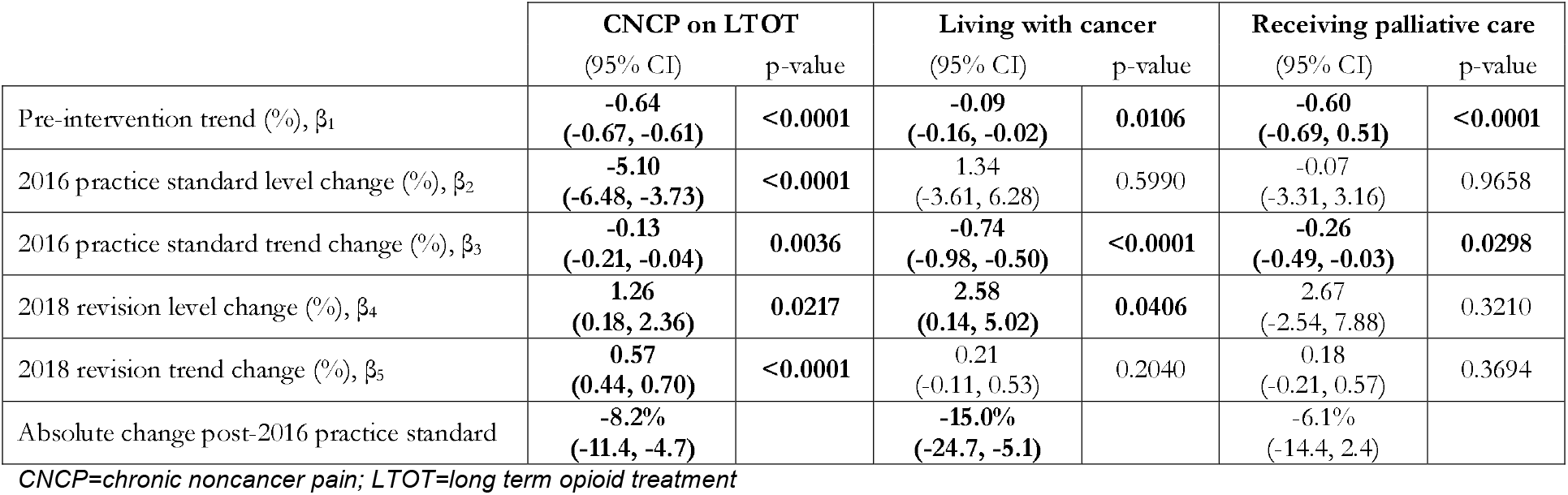
Changes in morphine milligram equivalents (MME) dispensed per person following the implementation of the 2016 practice standard and the 2018 revision, by pain type.

|  | **CNCP on LTOT** |  | **Living with cancer** |  | **Receiving palliative care** |  |
| --- | --- | --- | --- | --- | --- | --- |
|  | (95% CI) | p-value | (95% CI) | p-value | (95% CI) | p-value |
| Pre-intervention trend (%), $\beta_1$ | **-0.64**<br>**(-0.67, -0.61)** | **<0.0001** | **-0.09**<br>**(-0.16, -0.02)** | **0.0106** | **-0.60**<br>**(-0.69, 0.51)** | **<0.0001** |
| 2016 practice standard level change (%), $\beta_2$ | **-5.10**<br>**(-6.48, -3.73)** | **<0.0001** | 1.34<br>(-3.61, 6.28) | 0.5990 | -0.07<br>(-3.31, 3.16) | 0.9658 |
| 2016 practice standard trend change (%), $\beta_3$ | **-0.13**<br>**(-0.21, -0.04)** | **0.0036** | **-0.74**<br>**(-0.98, -0.50)** | **<0.0001** | **-0.26**<br>**(-0.49, -0.03)** | **0.0298** |
| 2018 revision level change (%), $\beta_4$ | **1.26**<br>**(0.18, 2.36)** | **0.0217** | **2.58**<br>**(0.14, 5.02)** | **0.0406** | 2.67<br>(-2.54, 7.88) | 0.3210 |
| 2018 revision trend change (%), $\beta_5$ | **0.57**<br>**(0.44, 0.70)** | **<0.0001** | 0.21<br>(-0.11, 0.53) | 0.2040 | 0.18<br>(-0.21, 0.57) | 0.3694 |
| Absolute change post-2016 practice standard | **-8.2%**<br>**(-11.4, -4.7)** |  | **-15.0%**<br>**(-24.7, -5.1)** |  | -6.1%<br>(-14.4, 2.4) |  |

*CNCP=chronic noncancer pain; LTOT=long term opioid treatment*

**Table 2.** Effects of 2016 practice standard and the 2018 revision on measures of care for patients on long-term opioid treatment (LTOT) for chronic non-cancer pain.

| Proportion of patients with: | No visit with prescribing physician within 90 days |  | Co-occurring benzodiazepine or other hypnotic |  | Supply exceeding 90 days or 250 pills |  | Daily dose exceeding 90 MME |  | Rapid taper |  |
| --- | --- | --- | --- | --- | --- | --- | --- | --- | --- | --- |
|  | (95% CI) | p-value | (95% CI) | p-value | (95% CI) | p-value | (95% CI) | p-value | (95% CI) | p-value |
| Pre-intervention trend (%), $\beta_1$ | -0.07<br>(-0.17, 0.04) | 0.2078 | **-0.67**<br>**(-0.71, -0.63)** | **<0.0001** | **-0.65**<br>**(-0.83, -0.47)** | **<0.0001** | **-0.53**<br>**(-0.57, -0.50)** | **<0.0001** | **-0.31**<br>**(-0.34, -0.28)** | **<0.0001** |
| 2016 practice standard level change (%), $\beta_2$ | -1.38<br>(-6.55, 3.79) | 0.5982 | **-5.24**<br>**(-7.67, -2.82)** | **<0.0001** | **-11.32**<br>**(-16.93, -5.72)** | **<0.0001** | **-3.56**<br>**(-4.66, -2.46)** | **<0.0001** | **1.85**<br>**(0.41, 3.29)** | **0.0128** |
| 2016 practice standard trend change (%), $\beta_3$ | -0.20<br>(-0.49, 0.09) | 0.1722 | **-0.62**<br>**(-0.74, -0.51)** | **<0.0001** | -0.24<br>(-0.60, 0.12) | 0.1935 | **-0.29**<br>**(-0.36, -0.22)** | **<0.0001** | **-0.13**<br>**(-0.21, -0.04)** | **0.0033** |
| 2018 revision level change (%), $\beta_4$ | 1.76<br>(-3.82, 7.33) | 0.5404 | **2.59**<br>**(0.25, 4.94)** | **0.0325** | 2.71<br>(-4.36, 9.77) | 0.4587 | -0.05<br>(-1.25, 1.35) | 0.9411 | 0.50<br>(-2.77, 3.78) | 0.7636 |
| 2018 revision trend change (%), $\beta_5$ | 0.22<br>(-0.25, 0.69) | 0.3558 | **0.74**<br>**(0.62, 0.86)** | **<0.0001** | **1.22**<br>**(0.77, 1.67)** | **<0.0001** | **0.48**<br>**(0.37, 0.59)** | **<0.0001** | -0.09<br>(-0.45, 0.27) | 0.6132 |
| Absolute change post-2016 practice standard | -6.1%<br>(-17.7, 6.0) |  | **-19.1%**<br>**(-24.0, -14.4)** |  | **-16.9%**<br>**(-30.4, -2.8)** |  | **-10.3%**<br>**(-13.0, -7.6)** |  | -1.2%<br>(-4.5, 2.3) |  |

**Figure 2.**
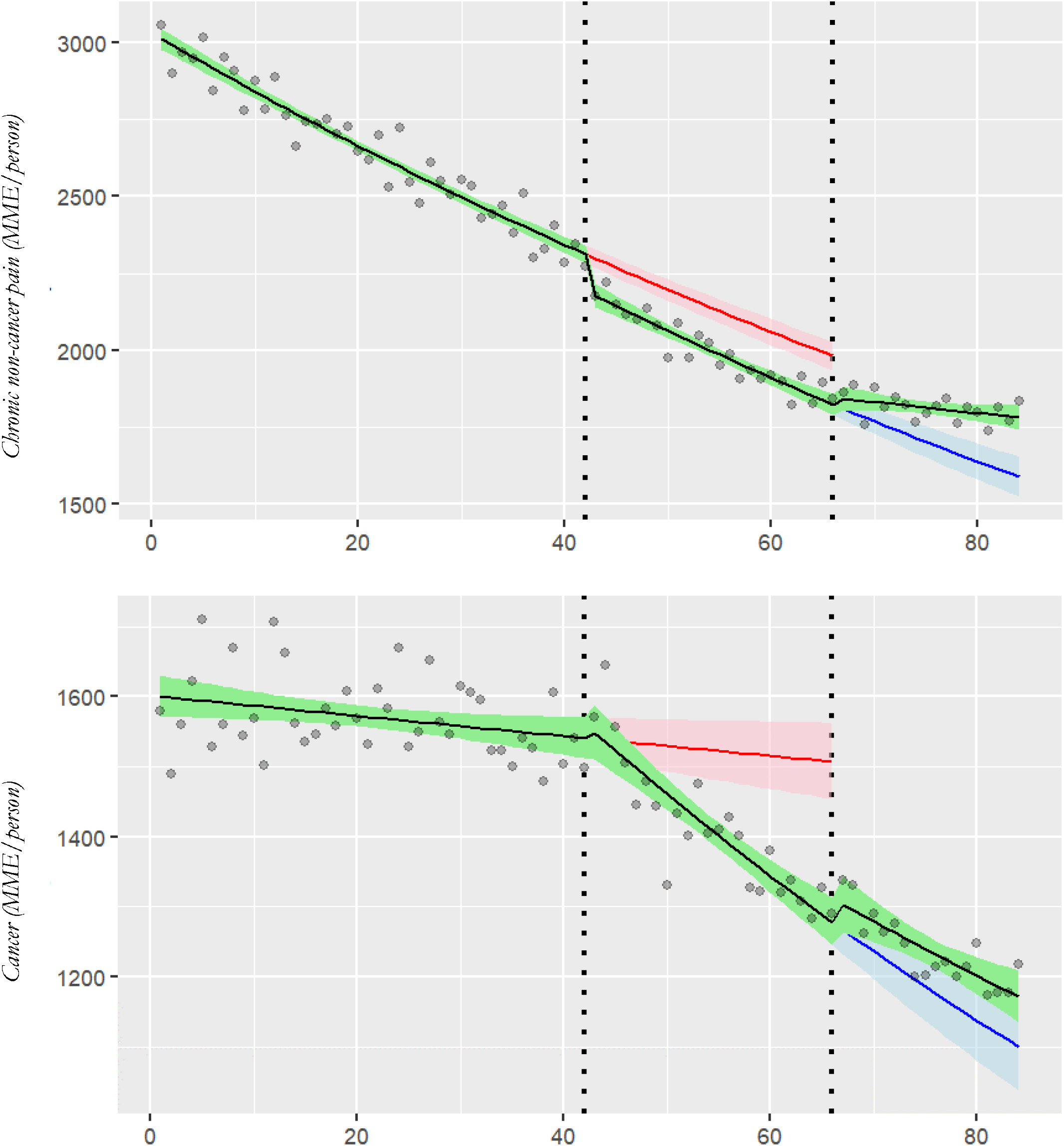
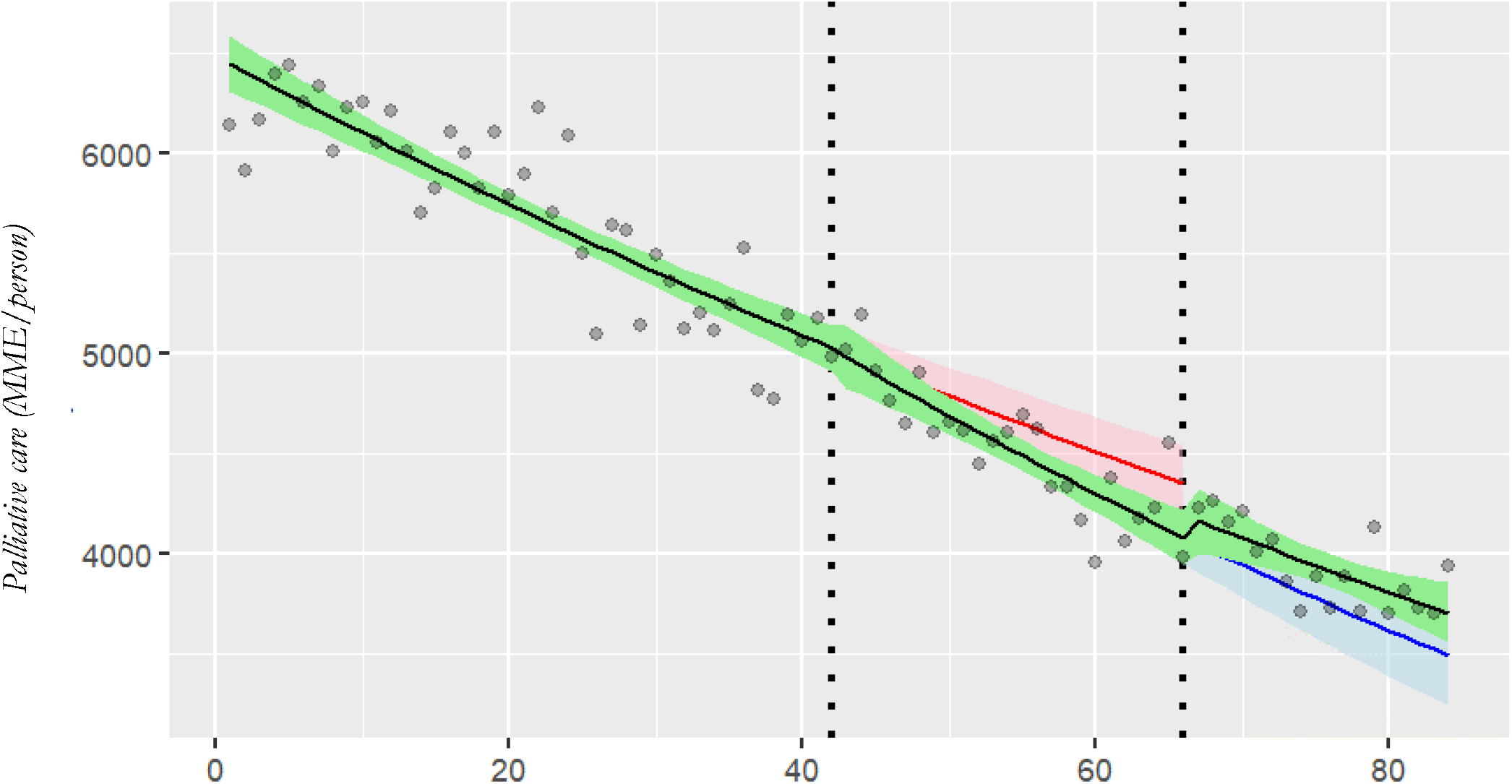
Changes in morphine milligram equivalents (MME) dispensed per person following the implementation of the 2016 practice standard and the 2018 revision, by pain type. The dashed lines at months 42 and 66 represent the implementation of the 2016 practice standard and the 2018 revision, respectively. Grey dots represent the total MME dispensed per person each month. Black solid line represents the trend in prescribing per period. The green ribbon represents the 95% CI around the observed trend lines. The solid red line represents the expected trend had the 2016 practice standard not been implemented, with the pink ribbon capturing the 95% CI. The solid blue line represents the expected trend had the 2018 revision not been implemented, with the light blue ribbon capturing the 95% CI.

**Figure 3.**
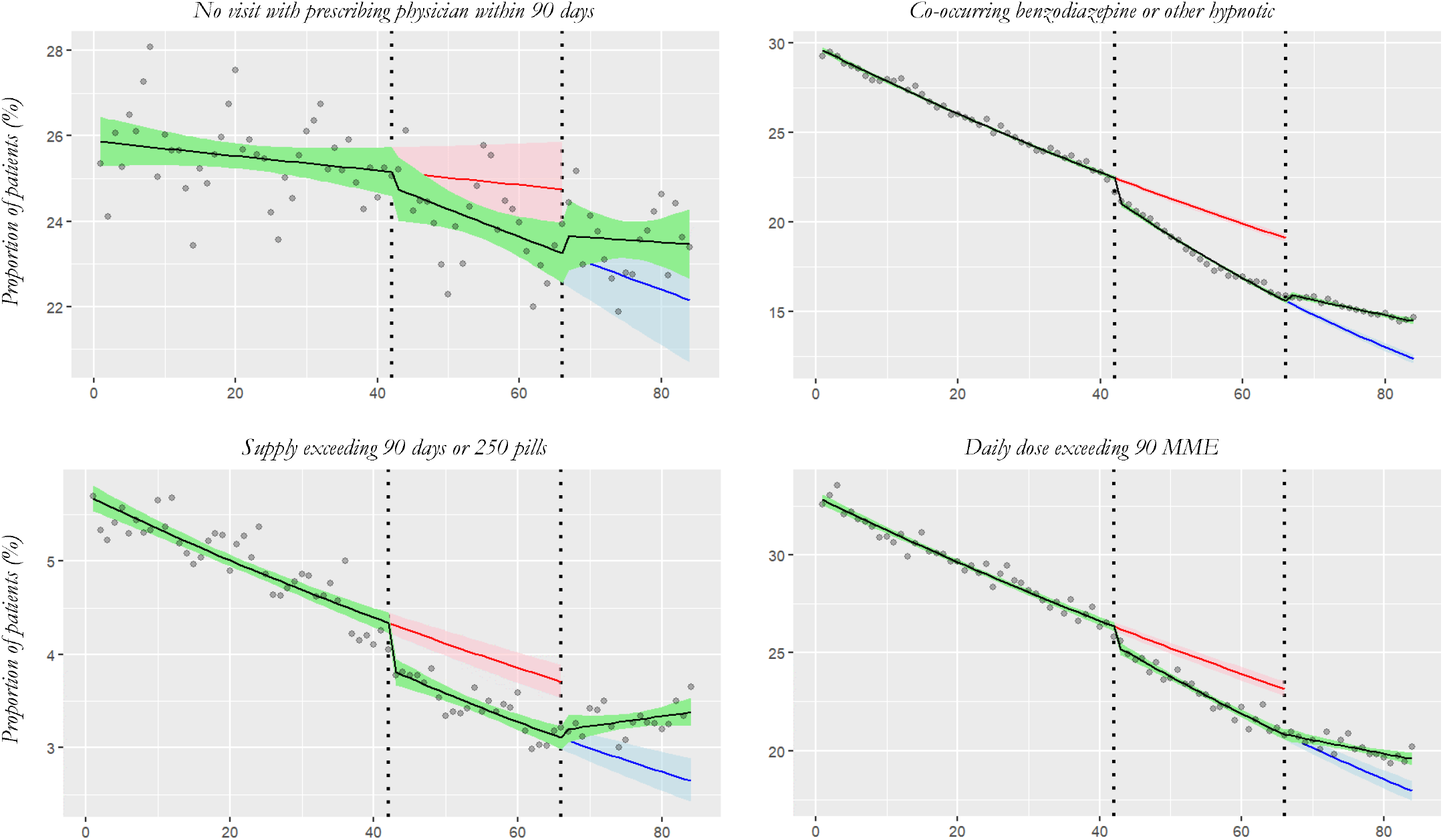
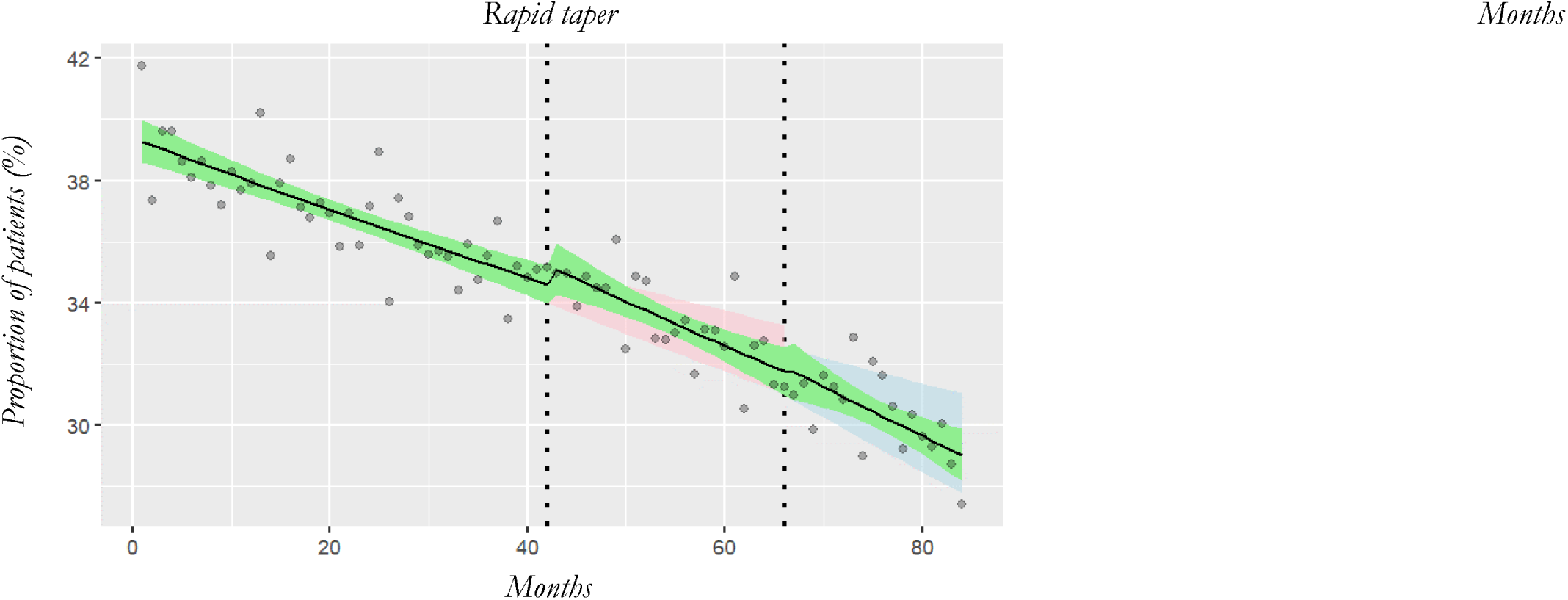
Effects of 2016 practice standard and the 2018 revision on proportion of patients on long-term opioid treatment (LTOT) for chronic noncancer pain with: a) no visit with prescribing physician within 90 days; b) co-occurring benzodiazepine or other hypnotic; c) supply exceeding 90 days or 250 pills; d) daily dose exceeding 90 MME; and e) rapid taper. The dashed lines at months 42 and 66 represent the implementation of the 2016 practice standard and the 2018 revision, respectively. Grey dots represent the monthly proportion as measured. Black solid line represents the observed trend in prescribing per period. The green ribbon represents the 95% CI around the trend lines. The solid red line represents the expected trend had the 2016 practice standard not been implemented, with the pink ribbon capturing the 95% CI. The solid blue line represents the expected trend had the 2018 revision not been implemented, with the light blue ribbon capturing the 95% CI.

a. with co-occurring benzodiazepine or other hypnotic dispensation (−0.6%, 95% CI: -0.7, - 0.5%);
b. with daily dose exceeding 90 MME (−0.3%, 95% CI: -0.4, -0.2%); and
c. rapidly tapered (−0.1%, 95% CI: -0.2, 0.0%).

The analysis also revealed that compared to the pre-intervention period, there was an immediate decline (level effect) in the MME dispensed per person (−5.1%, 95% CI: -6.5, -3.7%); and the proportion of patients with a co-occurring benzodiazepine or other hypnotic prescription (−5.2%, 95% CI: -7.6, -2.8%), whose supply exceeded 90 days or 250 pills (−11.3%, 95% CI: -16.9, -5.7%), and whose daily dose exceeded 90 MME (−3.6%, 95% CI: -4.7, -2.5%). However, we also observed a 1.9% (95% CI: 0.4, 3.3%) increase in the proportion of patients rapidly tapered. We observed no level or trend effects on the proportion of patients who did not have a visit to the prescribing physicians within 90 days. Meanwhile, the 2016 practice standard also decreased prescribing trends for the MME dispensed per person living with cancer (−0.7%, 95% CI: -1.0, -0.5%) and receiving palliative care (−0.3%, 95% CI: -0.5, 0.0%).

When the College released the June 2018 revision, we observed an increase (level effect) in the MME dispensed per person with CNCP (1.3%, 95% CI: 0.2 – 2.4%), and the proportion of patients with concurrent prescriptions for benzodiazepine or other hypnotics (2.6%, 95% CI: 0.3 – 4.9%). We also observed increases (trend effects) in the MME dispensed per person (0.6%, 95% CI: 0.4, 0.7%), and the proportion of patients with co-occurring benzodiazepine or other hypnotics 0.7% (95% CI: 0.6 – 0.9%), a supply exceeding 90 days or 250 pills (1.2%, 95% CI: 0.8 – 1.7%), and a daily dose greater than 90 MME (0.5%, 95% CI: 0.4 – 0.6%). For patients living with cancer, the MME dispensed increased 2.6% (95% CI: 0.1, 5.0%), while we observed no effect on opioids dispensed to patients receiving palliative care.

In absolute terms, twenty-four months after the 2016 practice standard’s implementation, MME dispensed per person were 8.2%, 15.0% and 6.1% lower than expected for patients treated for CNCP, living with cancer, and receiving palliative care, respectively.

## INTERPRETATION

Our analyses revealed that the 2016 practice standard influenced physicians’ opioid prescribing. Pre-existing declines in total opioid volume dispensed, high dose prescribing, co-prescribing with benzodiazepines or other hypnotics, supplies over 90 days or 250 pills, or daily doses above 90 MME accelerated. We also observed an immediate drop in MME dispensed per person, and the proportion of patients co-prescribed benzodiazepines or other hypnotics, receiving supplies over 90 days or 250 pills, and whose daily dose exceeded 90 MME; along with an increase in the proportion of patients rapidly tapered. When the 2016 practice standard was replaced in 2018, the downward trends slowed. For MME dispensed per person and proportion of patients with more than 90 days or 250 pills supply, declines reversed. Cancer and palliative care patients experienced the same prescribing trends.

Overall, our study findings align with descriptive papers noting declines in defined daily doses dispensed following the release of Canadian guidelines in 2017.(31, 32) Our findings are also consistent with US studies that observed declines in opioid prescribing,(14) increases in rapid tapering,(33, 34) and declines in the number of people treated with LTOT(35) following the implementation of the CDC’s 2016 prescribing guidelines. While our results appear to conflict with Crabtree et al.’s (2019) observation that the practice standard had no effect on opioid prescribing trends, two methodological differences between our work and theirs may explain this discrepancy. First, Crabtree et al.’s observation period may have been too short to detect the small but accumulating changes in opioid prescribing trends. Second, while we observed declines in MME dispensed per person for all three pain types, when aggregated across all patients, the total MME dispensed did not change dramatically. This is because for much of the observation period, the number of people living with cancer or receiving palliative care increased.

Together, these findings demonstrate that prescribing guidelines and practice standards can have immediate and long-lasting effects on physician prescribing. While most of the changes may be positive (e.g., fewer opioids in the community, a reduction in co-prescribed benzodiazepine etc.), incorrect interpretation can increase harms for some patients. Rapid tapering can have downstream consequences including people resorting to unregulated opioids for pain relief despite their risks.(36-38) Further, although practice standards may be specific to a pain type – they may have a chilling effect on access to opioids for non-target patient groups as well.

Our study has limitations that merit discussion. Since we used secondary administrative data and coding algorithms to define our cohort of people living with CNCP, we may have excluded people treated with opioids for intermittent/acute episodes of breakthrough pain (e.g., sickle cell crises). By prioritizing a conservative algorithm and using complete records of opioid dispensations in the community (e.g., irrespective of patients’ insurance coverage), we were able to examine the effects of the practice standard on the target population: primary care physicians responsible for most of the CNCP management in the community. Furthermore, we did not account for the potential effects of the 2017 *Canadian Guideline for Opioids Therapy and Chronic Non-Cancer Pain*, potentially attributing the combined effects of two prescribing interventions exclusively to the 2016 practice standard. Since the 2017 guideline was not legally enforceable and had softer language than the 2016 practice standard, the effects of this guideline on physician prescribing were expected to be small. Sensitivity analysis including a break for May 2017 (not shown here), did not reveal measurable effects. Finally, we only captured opioids dispensed from community pharmacies (i.e., excluded patients in long-term care). While increases in institutional dispensations may have offset observed trends, this is beyond this paper’s scope.(39)

## CONCLUSION

Our study found pre-existing declines in physicians’ opioid prescribing accelerated after the 2016 practice standard was implemented. This demonstrates the ability of practice standards to modify physician behaviour but also highlights how misinterpretation can harm patients. We recommend consulting patient groups affected by standards (or guidelines) before their release to reduce unintended consequences. Meanwhile, studies should measure spillover effects on exempt populations (e.g., changed opioid access for cancer or palliative care patients), and those in long-term care facilities. Colleges and government agencies should carefully assess the potential impacts of adopting practice standards.

## Supporting information

DINs used to exclude opioids prescribed for Cough, Diarrhea or Opioid Agonist Treatment

## Data Availability

All data produced in the present study are available upon reasonable request to the authors.

