## Supplementary material for "The effects of a provincial opioid prescribing standard on opioid prescribing for pain: interrupted series analysis": DINs used to exclude opioids prescribed for Cough, Diarrhea or Opioid Agonist Treatment

Appendix Table 1. Drug dispensation record using drug identification numbers (DIN)/product information number (PIN)

| Opioid agonist treatment | 00999792, 00999793, 02146126, 02242962 – 02242964, 02244290, 02295695, 02295709, 02394596, 02394618, 02408090, 02408104, 02424851, 02424878, 02453908, 02453916, 02468085, 02468093, 02474921, 02481979, 02483084, 02483092, 22123340, 22123346 – 22123349, 66123367, 66999990 – 67000020,  09850619, 09857499, 09857539 |
| --- | --- |
| Cough | 00003247, 00018694, 00023663, 00068594, 00068608, 00068756, 00093149, 00472549, 00507407, 00535230, 00550477, 00593168, 00593451, 00604658, 00690074, 00694827, 00779466, 01916564, 01916580, 01916599, 01916963, 01916971, 01934740, 01938363, 02049473, 02049481, 02053403, 02099748, 02169126, 02172917, 02198630, 02224577, 02230769, 02243063, 02244010, 02244011, 02244078, 02244079, 02244080, 02245592, 02245709, 02258099, 02324253 |
| Diarrhea | 00036323, 00346756, 00399345, 01909223, 02041634 |
